# An ultra-sensitive, ultra-fast whole blood monocyte CD169 assay for COVID-19 screening

**DOI:** 10.1101/2020.10.22.20215749

**Authors:** Moïse Michel, Fabrice Malergue, Inès Ait Belkacem, Pénélope Bourgoin, Pierre-Emmanuel Morange, Isabelle Arnoux, Tewfik Miloud, Matthieu Million, Hervé Tissot-Dupont, Jean-Louis Mege, Jean-Marc Busnel, Joana Vitte

## Abstract

CoVID-19 is an unprecedented epidemic, globally challenging health systems, societies, and economy. Its diagnosis relies on molecular methods, with drawbacks revealed by current use as mass screening. Monocyte CD169 upregulation has been reported as a marker of viral infections, we evaluated a flow cytometry three-color rapid assay of whole blood monocyte CD169 for CoVID-19 screening.

Outpatients (n=177) with confirmed CoVID-19 infection, comprising 80 early-stage (≤14 days after symptom onset), 71 late-stage (≥15 days), and 26 asymptomatic patients received whole blood CD169 testing in parallel with SARS-CoV-2 RT-PCR. Upregulation of monocyte CD169 without polymorphonuclear neutrophil CD64 changes was the primary endpoint. Sensitivity was 98% and 100% in early-stage and asymptomatic patients respectively, specificity was 50% and 84%. Rapid whole blood monocyte CD169 evaluation was highly sensitive when compared with RT-PCR, especially in early-stage, asymptomatic patients whose RT-PCR tests were not yet positive.

Diagnostic accuracy, easy finger prick sampling and minimal time-to-result (15-30 minutes) rank whole blood monocyte CD169 upregulation as a potential screening and diagnostic support for CoVID-19. Secondary endpoints were neutrophil CD64 upregulation as a marker of bacterial infections and monocyte HLA-DR downregulation as a surrogate of immune fitness, both assisting with adequate and rapid management of non-CoVID cases.

## Introduction

Severe acute respiratory syndrome coronavirus 2 (SARS-CoV-2), causing coronavirus disease 2019 (CoVID-19) is a historic global epidemic that continues to spread one year after having emerged (1). Its control requires targeted protection, distancing, and isolation. Aiming at selective isolation of subjects with a confirmed infection, a policy of massive diagnostic testing has been put in place in many countries. Ideally, this should allow any person to receive a prompt and straightforward answer at the time of symptom onset or contact tracing. This diagnosis is essentially based on molecular (reverse transcriptase-polymerase chain reaction (RT-PCR)) tests that detect viral RNA in a sample taken from the back of the nose or throat. Despite being the gold standard, RT-PCR has limitations. Firstly, the sensitivity is not optimal (2), mainly due to sampling quality and to the delay between contamination and colonization of the upper ear, nose, and throat area. Patients may receive a false negative result putting those around them at risk. Secondly, although RT-PCR is a fairly fast technique (1 to a few hours), laboratory saturation due to massive screening policies is leading to significant delays in sampling, processing, and result delivery, which may directly jeopardize distancing policies or unnecessarily hinder social and professional activity. Thirdly, deep nasal swabbing is unpleasant for the patient and potentially dangerous for the sampler. Finally, RT-PCR remains quite expensive. As a result and while massive testing is a required pillar for the fight against the pandemic, considerable efforts are made in search of efficient diagnostic aid solutions for mass testing strategies.

Antigenic tests of viral proteins are cheaper than RT-PCR, faster, and some can be performed on salivary samples that are easier to access and allow testing at home. However, their sensitivity is low, especially when using saliva instead of a nasal swab (3, 4). On the other hand, serological tests do not inform on actual carriage of the virus, are positive at least one week post-infection and lack sensitivity (5). In this context, harnessing immune markers of leukocyte activation is a promising tool. Indeed, leukocytes detect and rapidly respond to infection with secreted and surface activation molecules. We and others have previously reported that acute viral infections induce the appearance of CD169 (Siglec-1, sialoadhesin) at the surface of blood monocytes (6, 7). Monocyte CD169 expression is upregulated by type 1 interferons (8), produced by locally attacked tissues, and is found in all circulating blood monocytes, allowing its detection in minimal blood volumes such as a drop of blood at the fingertip. CD169 upregulation has been found in patients with HIV (9), EBV (10), RSV (11), CMV (12), Dengue (13, 14), Zika (15), noroviruses (16), Lassa and Marburg (17). Transcriptomic and mass cytometry studies have identified CD169 as a relevant biomarker for CoVID-19 (18, 19). The first evaluation by flow cytometry on patients not only confirmed CD169 as a SARS-CoV-2 infection marker, but also showed that its expression was much higher than for any other virus tested so far (20, 21).

Having developed a rapid (15 min) and affordable assay to measure monocyte CD169 upregulation in a few microliters of blood (22), we set out to assess its diagnostic efficacy in a large cohort of CoVID-19-confirmed patients, with SARS-CoV-2 RT-PCR as the reference method. This assay evaluated in parallel two other immune markers: upregulation of CD64 on polymorphonuclear neutrophils, which is widely used as an indicator of bacterial infection (23), and expression of HLA-DR on monocytes, which reflects the general state of the immune system (24): increased when activated by a pathogen (viral or bacterial), and decreased if the immune system is “exhausted” by a severe infection (e.g. sepsis).

## Results and Discussion

On the basis of the clinical and laboratory results, the 177 RT-PCR-confirmed SARS-CoV-2 patients were sorted into 3 groups: patients who presented at an early stage of the disease (less than 14 days after symptoms onset, n=80, group 1), those at a later stage of the disease (with a median of 19 days after symptoms onset (range 14-48), n=71, group 2), and asymptomatic patients (n=26, group 3). In each group, patients were further separated according to the concomitant RT-PCR results (**Figure 1**).

**Figure 1.**
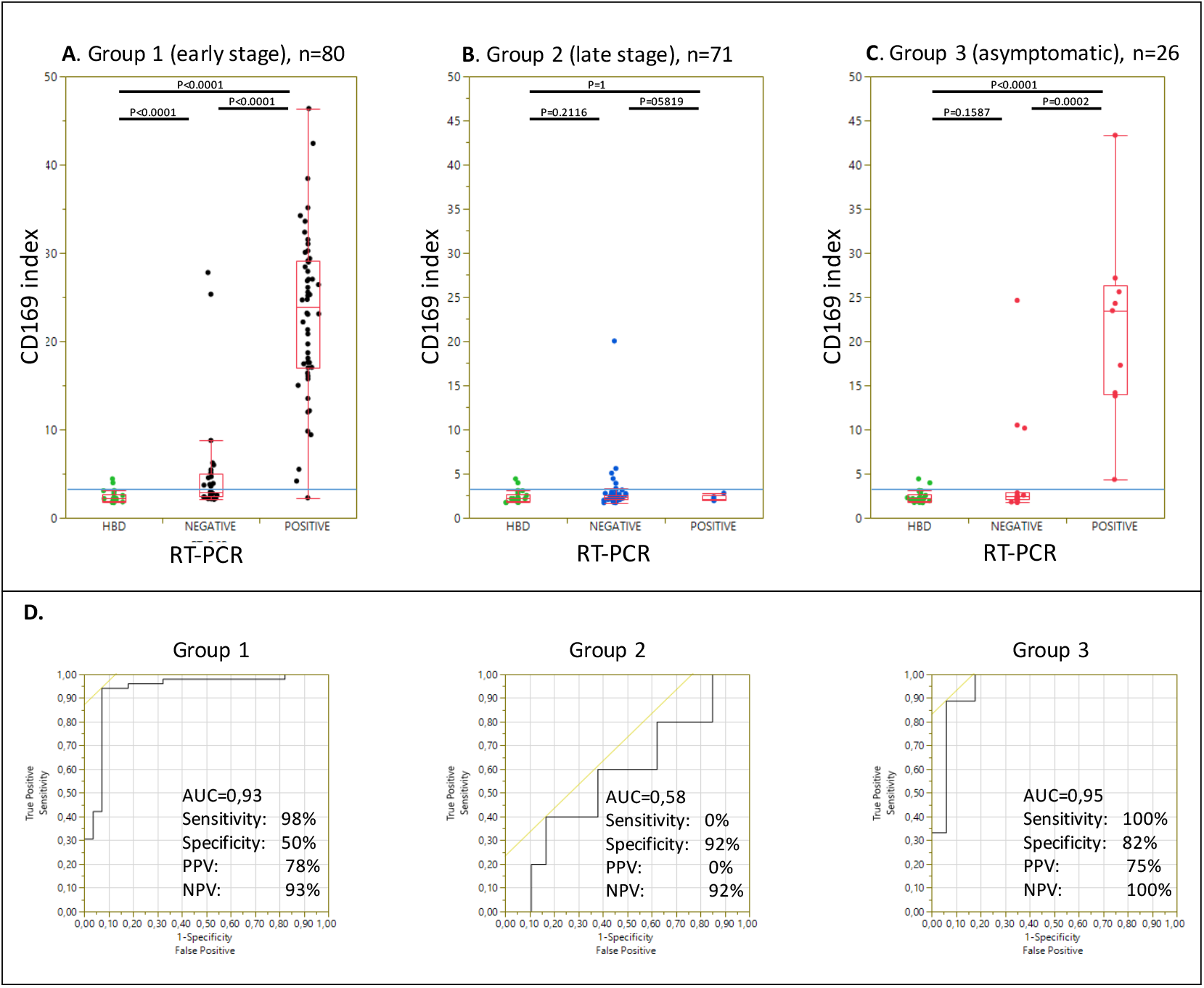
Expression of monocyte CD169 in CoVID-19 patients according to disease stage and SARS-CoV-2 RT-PCR results, compared to healthy blood donors. Box plots summarizing the level of CD169 index (ratio of monocyte vs lymphocyte signals) in Healthy Blood Donors (HBD, n=25, green dots) vs CoVID-19 patients at an early stage (A, black dots), late stage (B, blue dots), or asymptomatic (C, red dots). The blue line indicates the positivity threshold (3.5). Box-and-whisker plots come from the first to the third quartile and are cut by the median; segments at the end are extreme values. These 3 CoVID-19 groups are further split according to the concomitant RT-PCR results. ROC curves are calculated for each (D), then sensitivity, specificity, PPV, and NPV at the 3.5 threshold value. *AUC: Area Under the Curve; HBD: healthy blood donors; PPV: positive predictive value; NPV: negative predictive value*

CD169 expression was significantly higher in groups 1 and 3 as compared to group 2, with a median index of 17 (range 2.1-46.0, interquartile range IQR 4-31) and 2.8 (range 1.7-43.0, IQR 2.3-18.8) versus 2.3 (range 1.7-20.0, IQR 2.1-2.6), p<0.001. Using the previously established threshold of 3.5 for CD169 index (20), we observed that 23/25 (92%) healthy controls samples obtained from the blood bank were negative, with 2 outliers.

In the first group of CoVID-19 patients (early stage), the CD169 index was higher than the 3.5 threshold in 80% of patients (64/80) (**Figure 1A)**, while concomitant RT-PCR detected the virus (new cases), or confirmed it (re-tested cases), in 65% of patients (52/80). Among the 16 CD169-negative early-stage patients, 15 also had a negative concomitant RT-PCR. Sensitivity was 98%, with one patient exhibited a CD169 index below the threshold but a positive nasopharyngeal RT-PCR in the swab. Review of laboratory data for this patient showed very low and decreasing RNA quantities (cycle threshold (Ct) at 34.5 and 33.5, respectively 24 and 48 hours earlier), suggesting a near complete viral clearance. In line with this observation, the 15 other CD169 negative samples had been collected 6 to 14 days after the onset of symptoms, and the corresponding RT-PCR were also negative. Thus, sensitivity of monocyte CD169 was 98% when compared to RT-PCR.

In the second group (late stage), CD169 and RT-PCR were positive in only 7% of patients (5/71), in which the RT-PCR were negative (**Figure 1B**), indicating that CD169 expression returns to baseline levels upon viral clearance.

In the third group (asymptomatic cases), significant CD169 upregulation was detected in 46% of patients (12/26), at levels similar to those of group 1 comprising recent infections **(Figure 1C)**, while concomitant RT-PCR was positive in 35% of patients (9/26). All nine PCR positive cases were detected unambiguously. It is remarkable that almost half of asymptomatic patients, who made up 15% of the study cohort, expressed CD169 at the same level as patients experiencing symptoms. Indeed, the area under the curve (AUC) and overall performances, as displayed in **Figure 1D**, were similar for groups 1 and 3 (AUC at 0.93 and 0.95; sensitivity at 98 and 100 %; specificity at 50 and 82 %; positive predictive value (PPV) at 78 and 75 %; negative predictive value (NPV) at 93 and 100%, respectively for group 1 and 3) while CD169 index did not display good performance for patient with late stage disease (AUC at 0.58).

These findings suggest that despite the absence of symptoms, a systemic response orchestrating infection control takes place, rather than a purely local control of the infection (resistant tissue and/or tissue immunity).

There was only a weak correlation between CD169 level and RT-PCR Ct **(Figure 2)**. Still, the few CD169 negative patients were found among the weakest RT-PCR Ct.

**Figure 2.**
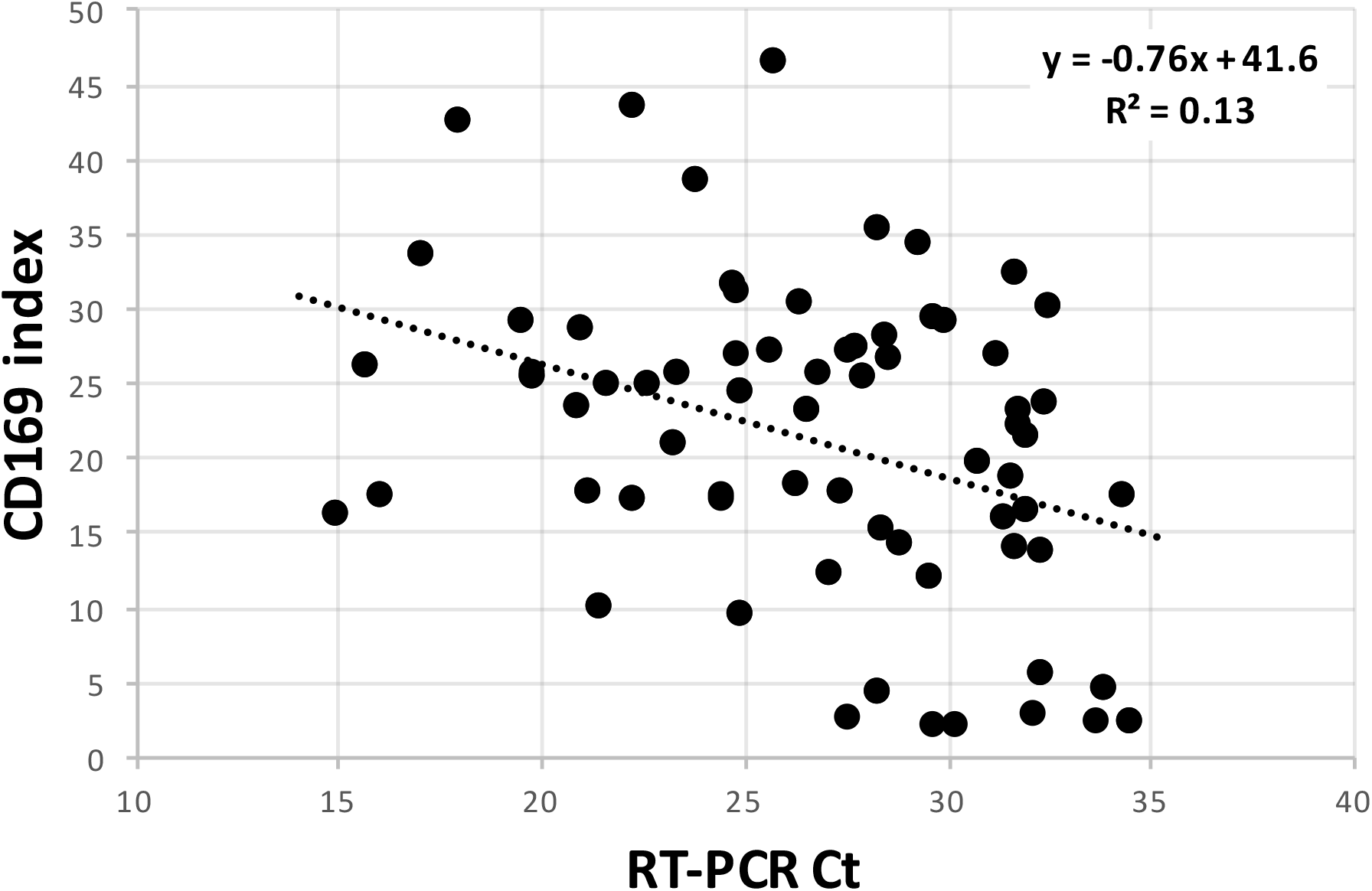
Correlation plot between concomitant SARS-CoV-2 RT-PCR Ct and CD169 index. Upregulation of monocyte CD169, expressed as CD169 index of monocyte-to-lymphocyte CD169 expression, was inversely correlated with SARS-CoV-2 Ct, itself inversely correlated with the patient’s viral load. *Ct, cycle threshold*.

Neutrophil CD64 expression, a marker of bacterial infections, was unchanged in 75% of the cases and weakly upregulated in 25% of the cases (45/177), showing no significant differences between groups (23, 27, and 31% respectively). Within this cohort of outpatients presenting with mild disease, HLA-DR was expressed at normal or slightly increased levels, an expected finding as opposed to the decrease usually observed in severe cases(25). **(Figure 3)**.

**Figure 3.**
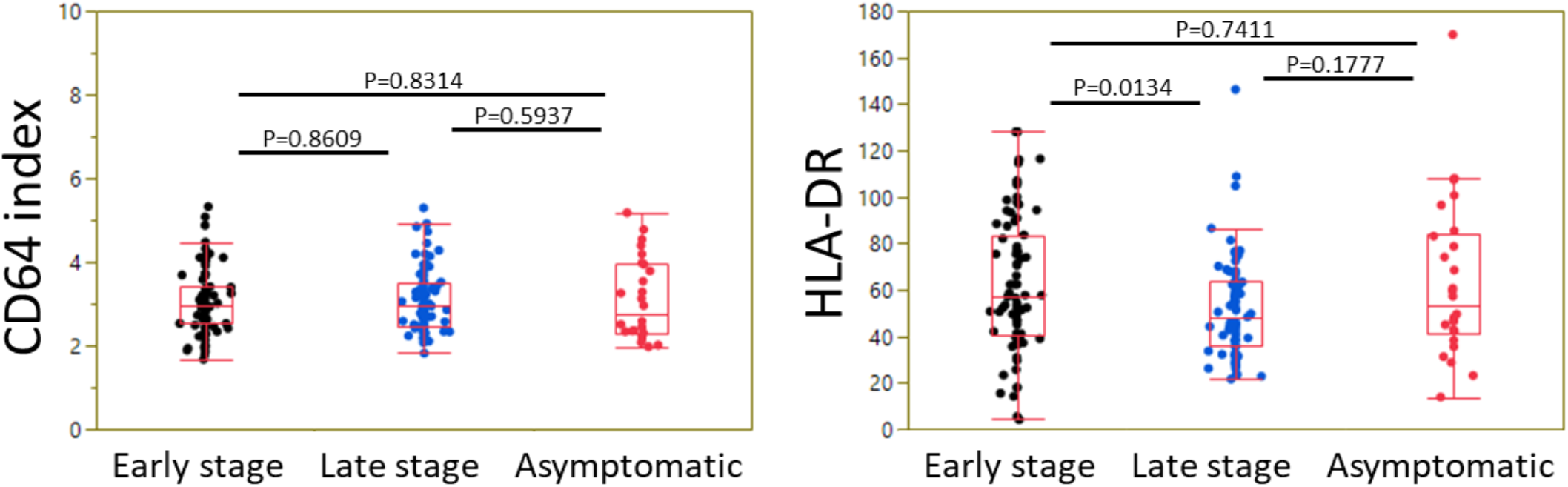
Expression of neutrophil CD64 and monocyte HLA-DR in CoVID-19 patients as a function of disease stage. Box plots summarizing the level of CD64 index (ratio of neutrophil vs lymphocyte signals) (A) and HLA-DR signal on monocytes (B) in CoVID-19 patients at an early stage, at a late stage, or asymptomatic, are shown. Box-and-whisker plots come from the first to the third quartile and are cut by the median; segments at the end are extreme values.

The sensitivity of monocyte CD169 upregulation was almost equivalent to RT-PCR for early-stage patients and asymptomatic patients. There was a high percentage (80%) of positive CD169 results in early-stage patients, peaking at 98.5% during the first week. As exemplified by these cases, false negative RT-PCR has a frequent occurrence, in agreement with the commonly described false negative rates ranging from 10 to 30%(5).

Considering the main screening target, *i*.*e*., recent symptomatic cases (within 7 days) with a positive RT-PCR the same day (n=49) or less than 48 h prior to or following CD169 assessment (n=54), 98.5% were identified through monocyte CD169 upregulation, with an unambiguous median index of 22.

Monocyte CD169 upregulation as a specific marker of viral infection was previously shown to outperform routine biomarkers (C-reactive protein, leukocyte counts, etc.). Indeed, CD169 elevation is restricted to acute viral infection(20). Virological identification of the culprit species, i.e. SARS-CoV-2 RT-PCR, remains necessary. Data obtained in the present study demonstrate that a testing strategy leveraging monocyte CD169 upregulation as a triage test could be designed. Taking into account the sensitivity of the assay and the current positivity rate of RT-PCR tests, ranging from 1 to 10%, a CD169-based screening could help prioritize true positives by a factor of 10 to 100, thereby opening the perspective to decrease the pressure currently observed on the health system.

Further advantages of monocyte CD169 assay are: 1. Better sensitivity that reduces or avoids false negative results. 2. Finger prick or venous blood sampling (easier, less painful, less dangerous, and without risk of failure). 3. A rapid result (15 minutes) allowing an immediate response to the consultation. 4. Affordable reagents and lighter logistics. 5. Not using a closed system, as the assay is supported by most flow cytometers equipping clinical laboratories.

Screening for monocyte CD169 upregulation could alleviate the load on specialized RT-PCR services and reduce overall costs, while making the sample collection step easier for the greatest number of people. One can even consider solutions where tests are performed at home and then sent to central laboratories by mail or carrier, since this antibody cocktail can be unitized and dried in tubes in a ready to use format stable at room temperature (Duraclone technology^®^, Beckman-Coulter Life Sciences).

From a more fundamental standpoint, considering the close relationship between type I interferon based signaling and monocyte CD169 expression, the latter marker could be employed as a surrogate for this cytokine family. If so, in the light of recent publications(26, 27) demonstrating the critical role of type I interferons in viral infections in general and in CoVID-19 in particular, rapid assessment of monocyte CD169 upregulation could help stratify patients and identify those at higher risk of developing more severe forms of COVID-19.

Combined detection of monocyte CD169 for viral infections, neutrophil CD64 for bacterial infections, and monocyte HLA-DR for immune status allows the referral of non-CoVID-19 cases to an adequate and rapid management. Moreover, the variety of neutrophil CD64 and monocyte HLA-DR levels observed in CoVID-19 patients may help identify patients at risk for developing more severe disease. This hypothesis would deserve a larger and longitudinal study.

The main strengths of this study are its outpatient design, its large sample size and the confirmation of SARS-CoV-2 infection with multiple RT-PCR. However, performing the comparison in real screening settings (finger pricks instead of venous blood) would provide further confirmation on the performance of the proposed assay. Prospective studies fulfilling this criterion and including further categories of patients (pediatric, comorbidities) are now required to fully assess the capabilities and possible limitations of this assay.

## Methods

### Samples

This non-interventional study was conducted in the immunology laboratory of the IHU Méditerranée Infection (Assistance Publique – Hôpitaux de Marseille, Marseille, France) on leftover samples from 177 consecutive patients aged 16 or older with RT-PCR-confirmed SARS-CoV-2 infection (at least one positive SARS-CoV-2 RT-PCR in nasopharyngeal swabs or tracheal aspiration).

Blood samples used in the flow cytometry study were obtained from patients being either first presentations, patients re-tested after a positive test in another laboratory, or positive patients re-tested during the clinical follow-up of the infection. Each patient was tested in parallel with a “concomitant RT-PCR” on the same day. Demographic, clinical and laboratory data including date of onset of SARS-CoV-2-related symptoms were collected for each patient retrospectively from electronical medical records. Samples of healthy blood donors (HBD) group served as controls (Convention N°7828, “Etablissement Français du Sang”, Marseille, France)

The study was approved by the institutional ethics and GDPR committee, with the reference number PJ4BQH. According to French law, the patients were informed and retained the right to oppose the use of their anonymized medical data for research purposes, but formal consent was not required for this non-interventional study.

### Flow cytometry

All antibodies and reagents were from Beckman-Coulter Life Sciences (Marseille, France). Leftover EDTA-anticoagulated samples were maintained at room temperature for a maximum of 24 hours prior to flow cytometry investigations. The 3 specific antibodies were pre-mixed in a ready-to-use cocktail (prototype of the IOTest Myeloid Activation CD169-PE/HLA-DR-APC/CD64-PacBlue Antibody Cocktail, Beckman Coulter Life Sciences). The cocktail was then pre-mixed with 0.5 mL of Versalyse RBC lysing solution, and 10 µL of blood were added in the reaction tube. The mixture was finally mixed manually. After 10 minutes incubation at room temperature, the samples were analyzed 1 minute on a routine 3 laser Navios flow cytometer from Beckman Coulter Life Sciences (Miami, USA).

### Data analysis and statistics

Flow cytometry data files were analyzed using the Kaluza software, version 2.1 (Beckman Coulter Life Sciences). Leukocytes were gated using Side Scatter (SSC) and CD64 expression as lymphocytes (low SSC CD64-), monocytes (intermediate SSC CD64+), and neutrophils (high SSC), prior to the analysis of CD169, CD64 and HLA-DR level of expression. The CD169 index was calculated as the ratio of CD169 expression on monocyte vs CD169 expression on lymphocyte (background) signal. Similarly, the CD64 index was calculated as the ratio of neutrophil versus lymphocyte signal. Data were exported to JMP 14.2.0 software (SAS) for statistical analysis. Mean index values of each group were compared via Student’s or Kruskal-Wallis tests as appropriate. A two-sided p-value < 0.05 was considered statistically significant.

## Data Availability

All data referred to in the manuscript are available

## Author contributions

JV, TM and PEM designed the study. MoM, IAB and FM conducted experiments and acquired data. MaM and HTD supervised clinical procedures. MoM collected and analyzed demographic, clinical and laboratory data. PB and FM provided antibody panels and PEM and IA the flow cytometry platform. MoM, FM, and JMB analyzed experimental data. FM, MoM, JMB, and JV wrote the manuscript. JLM revised the manuscript for important intellectual content. All authors read and approved the final manuscript.

---

**Table 1.** Demography of the study cohort. Early stage is defined as the 14 first days from symptom onset; late stage is defined as 15 days or later after symptom onset. Data are presented as median and 5th-95 th percentiles. *NA, not applicable; Nb: Number*

|  | Early stage | Late stage | Asymptomatic | Total | p-value |
| --- | --- | --- | --- | --- | --- |
| **n** | 80 | 71 | 26 | 177 |  |
| **Age** | 45.8<br>(19.4 – 81.9) | 45.0<br>(22.3 – 68.7) | 45.0<br>(20.5 – 61.3) | 45.3<br>(20.5 – 69.6) | 0.98 |
| **M/F gender ratio** | 0.82 | 0.61 | 1.89 | 0.82 | 0.056 |
| **Time from symptom onset** | 6<br>(2 – 14) | 20<br>(15 – 37) | NA | 16<br>(3 – 35) | $10^{-17}$ |
| **Nb of SARS-CoV-2 RT-PCR positive** | 52<br>(65 %) | 5<br>(7.04 %) | 9<br>(34.62 %) | 76<br>(42.94 %) | $10^{-12}$ |
| **SARS-CoV-2 RT-PCR Ct value** | 27.25<br>(17.60 – 32.39) | 30.20<br>(28.02 – 33.38) | 27.70<br>(18.58 – 33.30) | 27.65<br>(17.33 – 33.40) | 0.21 |

